# Effects of Joint Mobilization on Pain, Range of Motion, and Function in Patients with Knee Osteoarthritis: A Systematic Review and Meta-analysis of Randomized Controlled Trials

**DOI:** 10.1101/2024.11.10.24317074

**Authors:** Xiaotin Chen, Debiao Yu, Yaoyu Lin, Peng Chen, Bin Shao, Fuchun Wu

## Abstract

**Background:** Knee osteoarthritis (KOA) is a common degenerative joint disease that severely affects patients’ quality of life. Joint-mobilization technique reportedly improves joint pain, limited mobility, and dysfunction significantly. This meta-analysis aimed to systematically assess the clinical efficacy of joint-mobilization technique treatment on the level of knee pain, function, and mobility in patients with KOA. We also aimed to provide evidence-based medical data for the clinical management of KOA.

**Methods:** We searched four English databases (PubMed, Web of Science, Embase, and Cochrane) and three Chinese databases (China Biomedical Literature Database, CNKI, VIP, and Wanfang). The search date was from the date of inception to February 1, 2024 for each database. Randomized controlled trials investigating the efficacy of joint release in KOA were identified. Meta-analysis was performed using RevMan 5.4 and Stata 17.0.

**Results:** A total of 8 studies involving 432 patients with KOA were included. Our meta-analysis showed that compared with the control group, the experimental group showed a significant improvement in knee pain level (SMD=-1.69, 95% CI [-1.74, -0.82] Z=3.96 P<0.0001), WOMAC scale (SMD=-0.74 95% CI [-1.39, -0.10] Z= 2.25 P=0.02) were significantly improved. However, they were not superior to controls in knee flexion (SMD=2.3 95% CI [0.98, 3.62] Z=3.41 P=0.00006) and extension mobility (SMD=1.79, 95% CI [1.38, 2.20], Z=8.54,P<0.00001).

**Conclusion:** Joint-mobilization technique has some advantages in improving the degree of knee pain and dysfunction in patients with KOA, but it is not better than the control group in improving knee mobility. This study provides theoretical support for the promotion of joint-mobilization technique in KOA treatment.

## 1 Introduction

Knee osteoarthritis (KOA) is a common degenerative osteoarthritic disease characterized by the narrowing of the joint space, destruction of articular cartilage, and formation of bony encumbrances^[1]^. An estimated 250 million people worldwide reportedly suffer from KOA, which primarily affects people over 45 years old and is more common in females^[2, 3]^. Indeed, it has become the fourth most disabling disease in the world^[4]^. Clinical manifestations include severe joint pain, stiffness, limited mobility, and dysfunction, and patients often seek medical attention due to severe joint pain and limited mobility^[5]^. Thus, the primary goal of treatment for KOA is to relieve joint pain, improve joint mobility, and improve the function of the knee joint, thereby improving the patient’s quality of life^[6]^. Current treatments for KOA are primarily non-surgical, such as using drugs, physical factor therapy, acupuncture, and so on^[7]^. However, it has long-term treatment, poor efficacy, or other problems in some patients^[8]^. Joint-mobilization technique is a manipulation to relieve joint pain and maintain or improve joint range of motion^[9]^. It is extensively used for joint dysfunction caused by any mechanical factor, including Maitland, Kaltenborn, and Mulligan dynamic joint mobilization^[10]^. Joint-mobilization techniques initiate local physiological mechanisms and also involve central mechanisms, such as promoting the activation of inhibitory pathways in the spinal cord or higher levels of downstream inhibitory pathways in the brainstem^[11]^.

Joint-mobilization technique is a manipulative therapy technique in which the therapist manipulates the joint through passive movements (physiological or accessory movements)^[10]^. Different amplitudes and speeds within the range of motion permitted by the joint are utilized to reduce or cure joint pain, increase proprioceptive feedback, and improve the range of motion of the joint^[12]^. The most commonly used types today include Maitland mobilization, Kaltenborn mobilization technique, and Mulligan mobilization with movement^[13-15]^. Among them, Maitland and Kaltenborn mobilization are graded according to the force applied by the therapist, with Maitland categorized as a grade 5 and Kaltenborn as a grade 3, thereby providing a degree of objectivity^[16]^. Mulligan mobilization with movement can be performed either actively by the patient or passively by the therapist through continuous gliding within the physiological movement of the joint^[17]^. Currently, joint-mobilization techniques are widely used for joint dysfunction caused by any mechanical factor^[18, 19]^. Clinical studies have shown that joint-mobilization techniques have significant therapeutic effects in other diseases, such as frozen shoulder and spondyloarthropathies^[20-22]^.

In a similar review paper, LI, et al^[23]^analyzed the effect of one type of joint-mobilization technique only on KOA, and no specific treatment suggestion is provided. Existing reviews have not performed a meta-analysis of pain, joint mobility, and function of all types of joint-mobilization techniques for KOA treatment. Thus, this study aimed to evaluate the efficacy of joint mobilization techniques in adherence to the Preferred Reporting Items for Systematic Reviews and Meta-Analyses (PRISMA) guidelines. We conducted a methodological evaluation of eligible randomized controlled trials and clinical effectiveness. A meta-analysis of the effectiveness outcomes of the joint-mobilization techniques was then conducted. The effects of the joint-mobilization intervention on pain, joint mobility, and functional outcomes in patients with KOA were determined.

## 2 Methods

This research is a systematic review and group analysis, mainly involves the study of previously published data review rather than direct contact with patients or for personal health information. Therefore, no new ethical approval or patient consent is required for this type of investigation according to international research ethics guidelines.We followed PRISMA guidelines for this study. The protocol was registered in PROSPERO (registration no. CRD42023481795)

### 2.1 Literature search

We searched seven databases, including four English databases (PubMed, Web of Science, Embase, and Cochrane) and three Chinese databases (China Biology Medicine disc, CNKI, VIP, and Wanfang). The search date was from the inception date of each database to February 1, 2024, and languages were Chinese and English. The search was performed using a combination of subject terms and free words. The following medical terms were used for the search: "osteoarthritis of the knee,” "joint mobilization," "mulligan,” "Maitland,” "Kaltenborn," and "Musculoskeletal Manipulations." The search strategy is shown in Table 1.

**Table 1.**
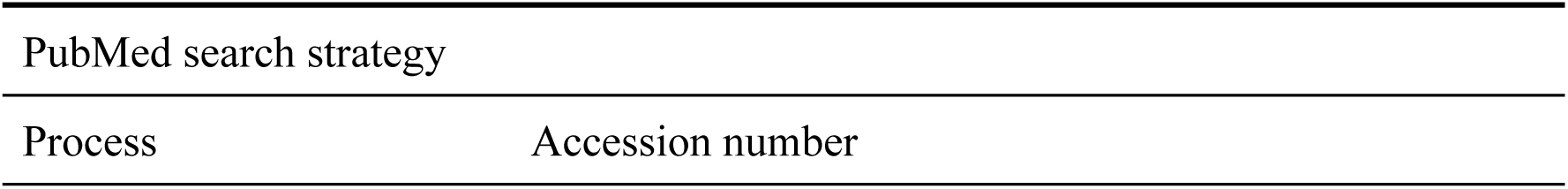
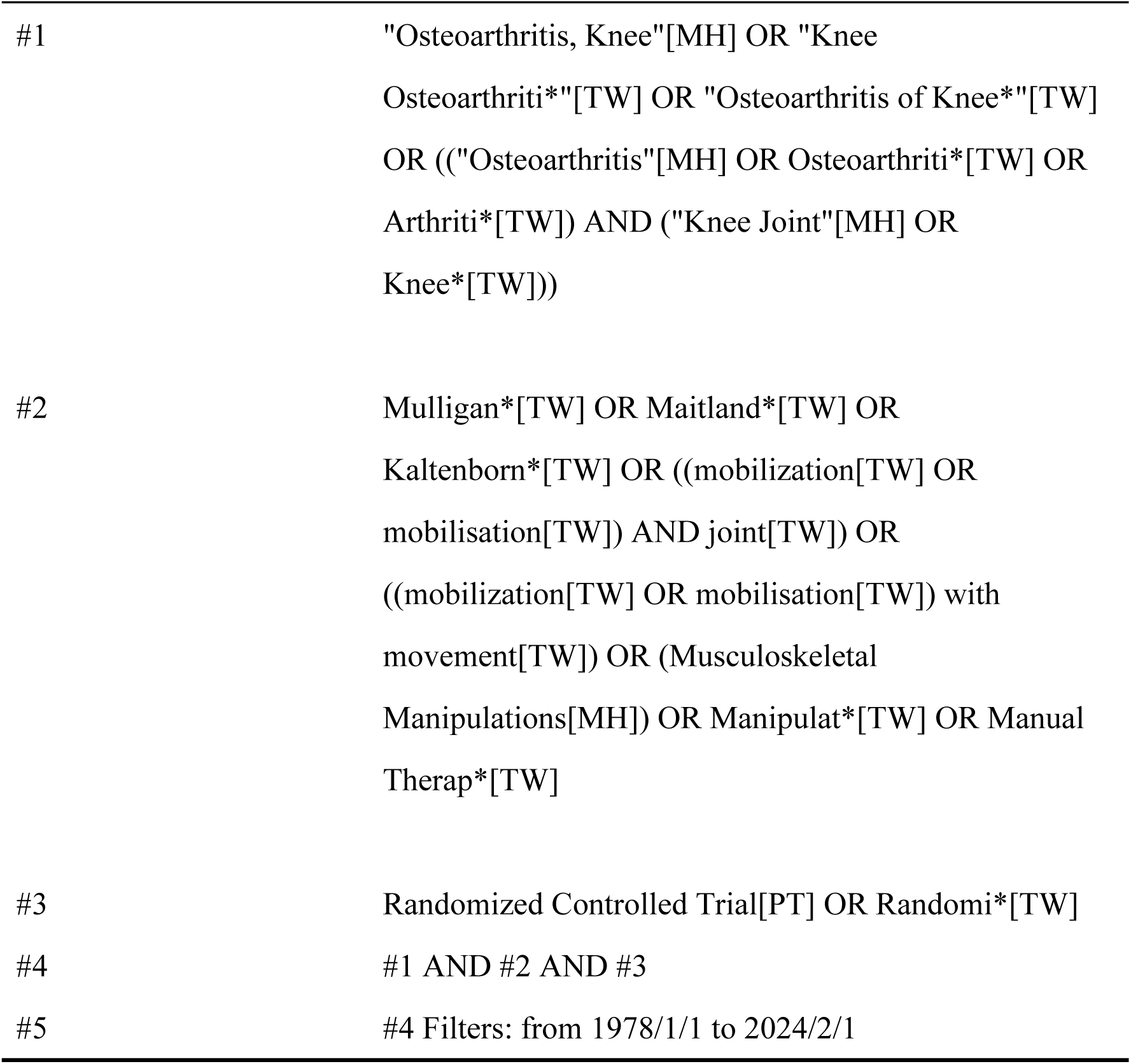
Search strategy for PubMed.

### 2.2 Literature inclusion criteria

The inclusion criteria were as follows. For study type, we included randomized controlled trials investigating the effects of joint-mobilization technique on knee pain, function, and mobility in patients with KOA limited to publications in Chinese and English.

For study participants, patients with osteoarthritis of the knee diagnosed by imaging of any age were included.

For intervention, we included studies in which the control group received sham control, waitlist treatment, standard medication, or other active interventions, and the experimental group received Maitland, mulligan, Kaltenborn, or other joint-mobilization techniques. The primary goal was to relieve pain, limited mobility, and dysfunction in patients with KOA. The duration of the intervention was limited to 2–6 weeks.

For outcome measures, a study was included if the Visual Analog Scale (VAS) was used to assess the patients’ knee pain level; knee function in patients with KOA was assessed using the Western Ontario and McMaster Universities Arthritis Index (WOMAC); and knee mobility was assessed using knee flexion and extension angles.

### 2.3 Literature exclusion criteria

The exclusion criteria were as follows: duplicate publications, as well as non-randomized controlled trials, including animal experiments, literature reviews, systematic reviews, meta-analyses, case reports, expert summaries, operational research, parametric studies, prospective or retrospective clinical observations, reviews, letters, conference abstracts, etc.; the contact authors were unable to obtain useful data; knee pain, limited mobility, and dysfunction due to other causes; and the observation group received a different method of combined treatment than the control group, in addition to joint-mobilization technique.

### 2.4 Literature screening

As per the search protocol, two separate researchers conducted searches across electronic databases and additional resources. Titles, abstracts, and full texts of articles underwent comprehensive evaluation. In cases of disagreement between the two researchers, the ultimate determination was reached through deliberation involving a third researcher.

### 2.5 Data extraction

Two separate researchers compiled data into Excel spreadsheets (Microsoft Corporation, Redmond, WA), which encompassed authors’ names, publication years, study designs, participant counts per group, specifications of experimental and control groups, treatment duration and frequency, as well as outcome assessments. In cases where discrepancies arose between the Excel spreadsheets of the two researchers, a third researcher adjudicated to arrive at a final decision.

### 2.6 Bias risk and quality assessment

The risk of bias was evaluated utilizing the Bias Risk tool devised by the Cochrane Collaboration. Biases were assessed across various domains, including random sequence generation, allocation concealment, blinding of participants and personnel, blinding of outcome assessment, incomplete outcome data, selective reporting, and other potential biases. Each domain underwent scrutiny by two independent researchers who categorized the risk into three levels: high, low, or unclear. In instances where there was a discrepancy in the assessment of bias between the two researchers, a consensus was reached through consultation with a third researcher, who then finalized the decision.

### 2.7 Statistical analysis

Statistical software RevMan 5.4 and Stata 17.0 were used for the meta-analysis of the above outcome indicators. For count data, relative risk served as the statistical measure, whereas for measured data, standardized mean difference (SMD) was used, and the significance of SMD was assessed by Z-test. All effect sizes were expressed as 95% CIs (CIs). Heterogeneity was assessed using the Q-test. When I^2^ ≤ 50%, a fixed-effect model was selected for analysis, whereas when I^2^ > 50%, a random-effect model was used. Sources of heterogeneity were explored through subgroup analysis or sensitivity analysis. If the cause of heterogeneity cannot be determined, descriptive analysis was conducted based on the original literature. Article publication bias analysis was performed using Egger’s test (performed in Stata 17.0 software), and *P* < 0.05 was considered a significant risk of publication bias. Meanwhile, *P* > 0.05 was considered a non-significant risk of publication bias, and conclusions were reliable.

## 3 Results

### 3.1 Literature search results

A preliminary search yielded 2923 articles, which were then organized using Endnote software. Following the removal of 832 duplicates, screening of titles and abstracts of the remaining studies ensued to eliminate reviews, study protocols, case reports, self-controlled studies, animal experiments, as well as subjects and means of intervention deemed irrelevant. Consequently, 1551 articles were excluded from further consideration. Subsequent meticulous examination of full-texts resulted in the exclusion of an additional 532 papers due to ambiguous diagnostic criteria, absence of outcome indicators, unclear treatment descriptions, or lack of data availability. Ultimately, 8 trials met the inclusion criteria for this study. The schematic representation of the literature screening process is depicted in Figure 1.

**Figure 1.**
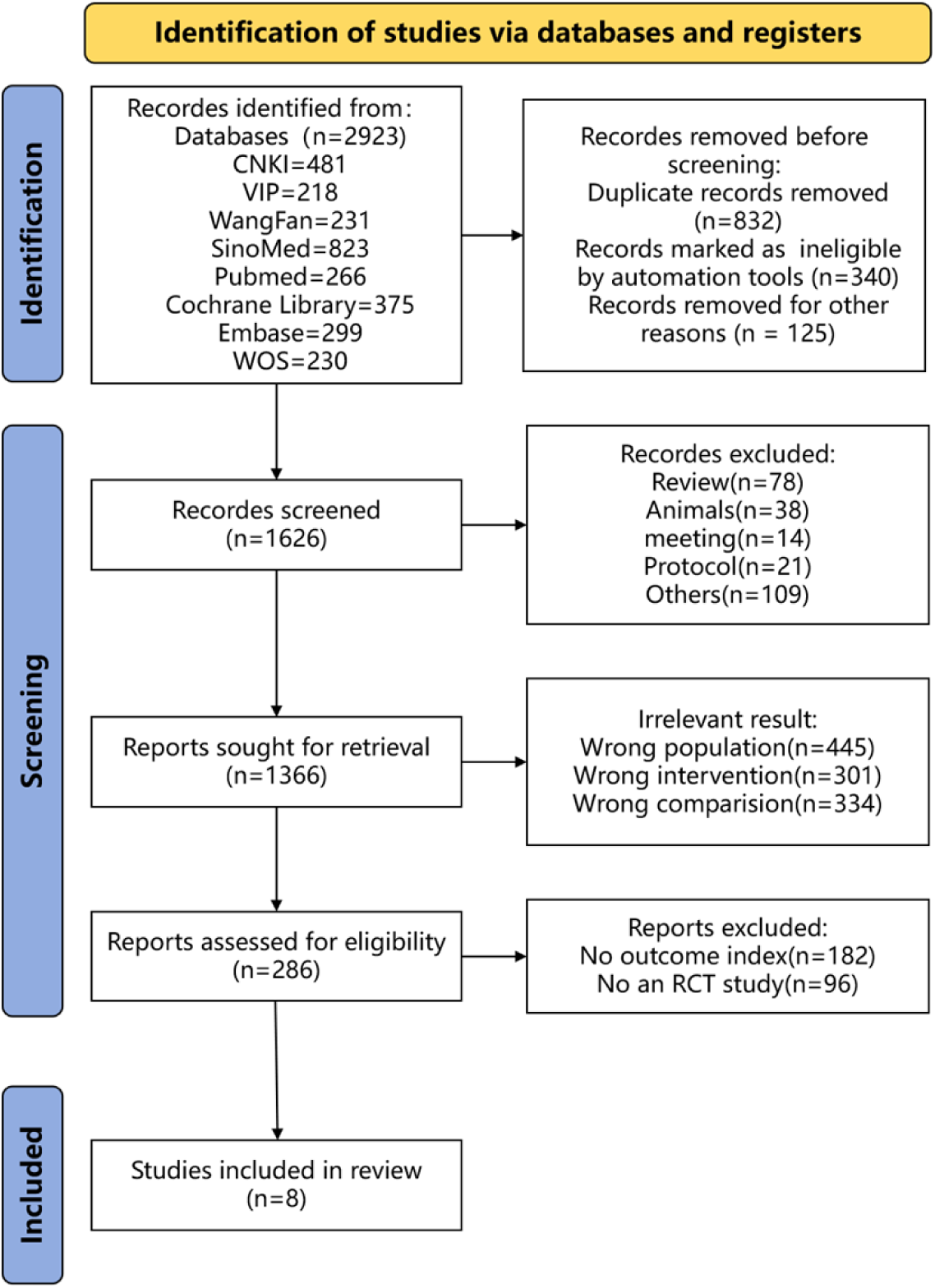
Literature screening process.

### 3.2 Characteristics of included literature

Eight trials were deemed eligible for inclusion in this meta-analysis, comprising two written in Chinese and six in English. The combined participant pool consisted of 432 individuals with knee osteoarthritis (KOA), with 215 allocated to the experimental group and 217 to the control group. The studies were published between 2007 and 2023. The experimental group used a combination of joint-mobilization techniques, including Mulligan, Maitland, joint traction, or patellar release with basic treatment for the control group. Moreover, this group received interventions such as pharmacological treatment physical factor therapy and manipulative massage. Among these studies, 6 reported pain scores^[24-29]^, WOMAC had 5^[24-26, 30, 31]^ and 3^[24, 26, 28]^ in knee flexion angles and 2^[24, 28]^ in knee-extension angles. The essential features of the literature included in this analysis are detailed in Table 2.

**Table 2.**
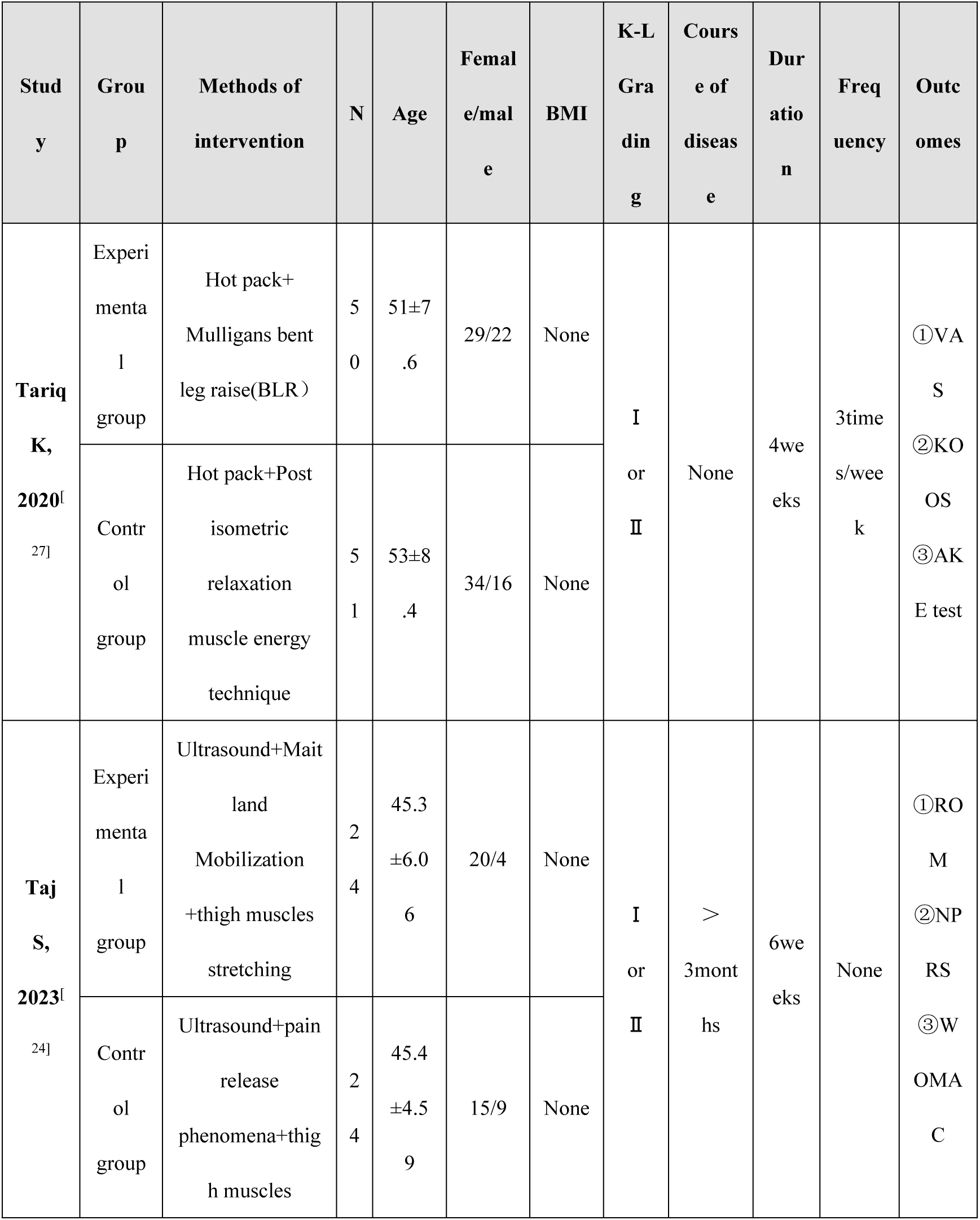
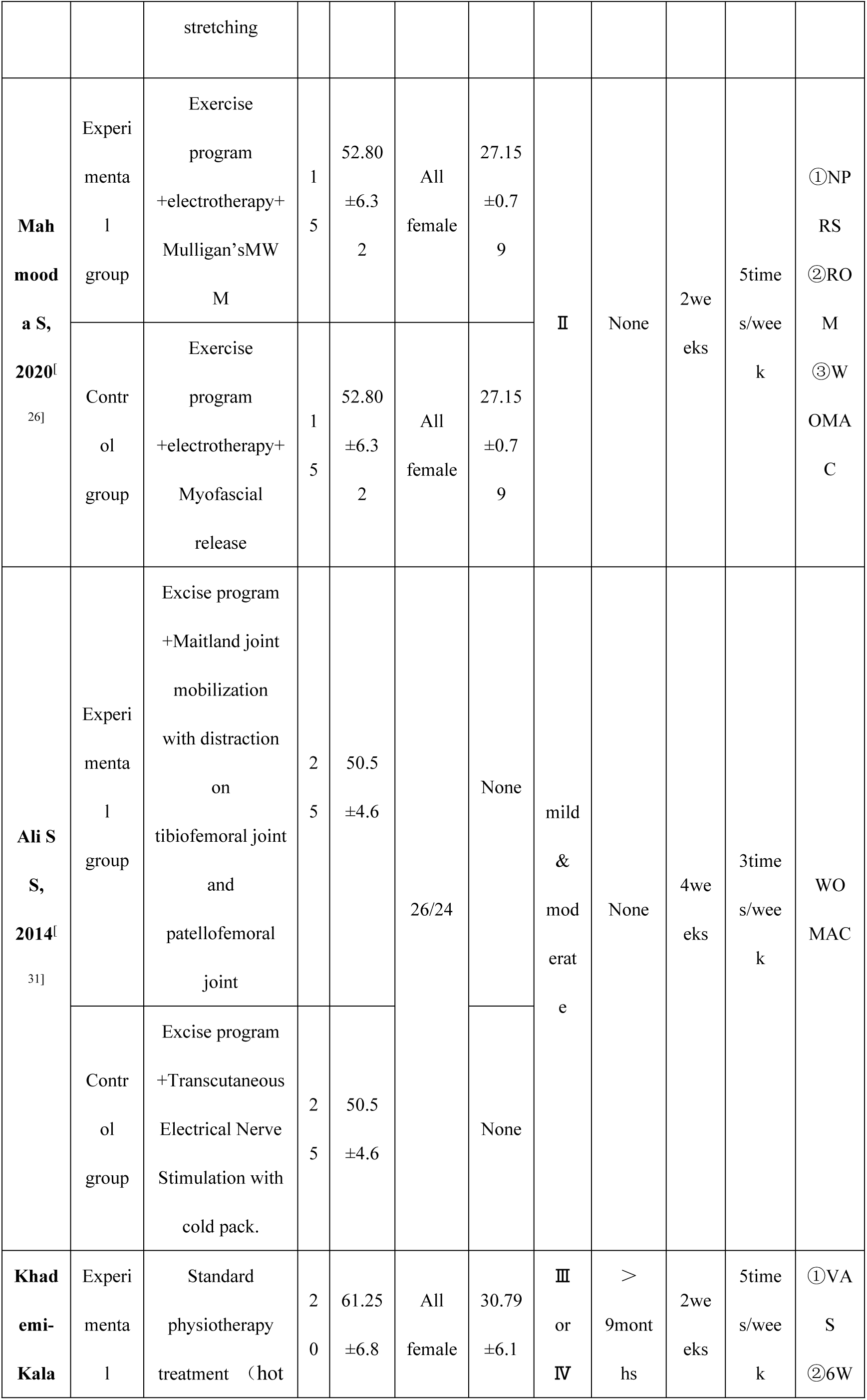
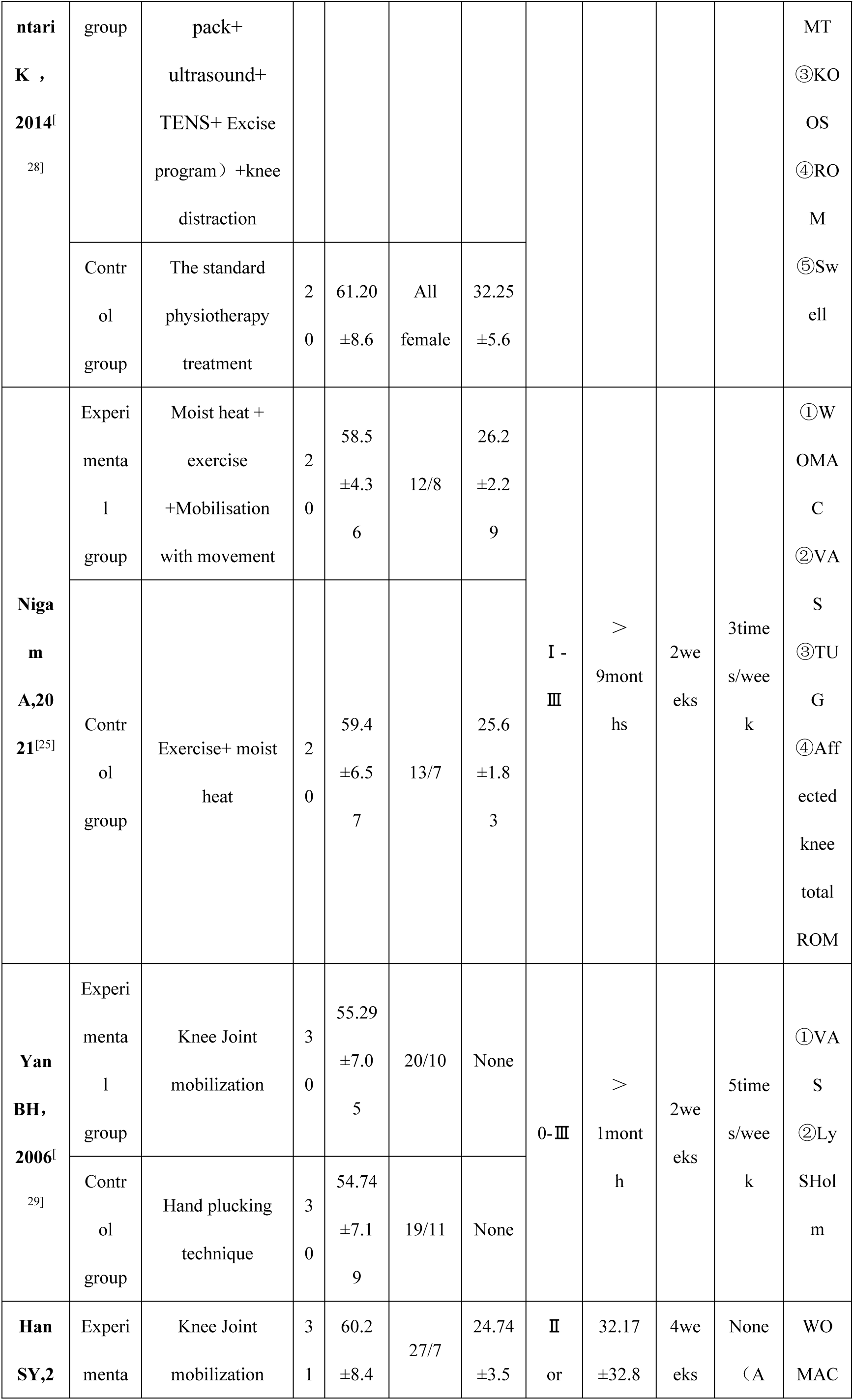
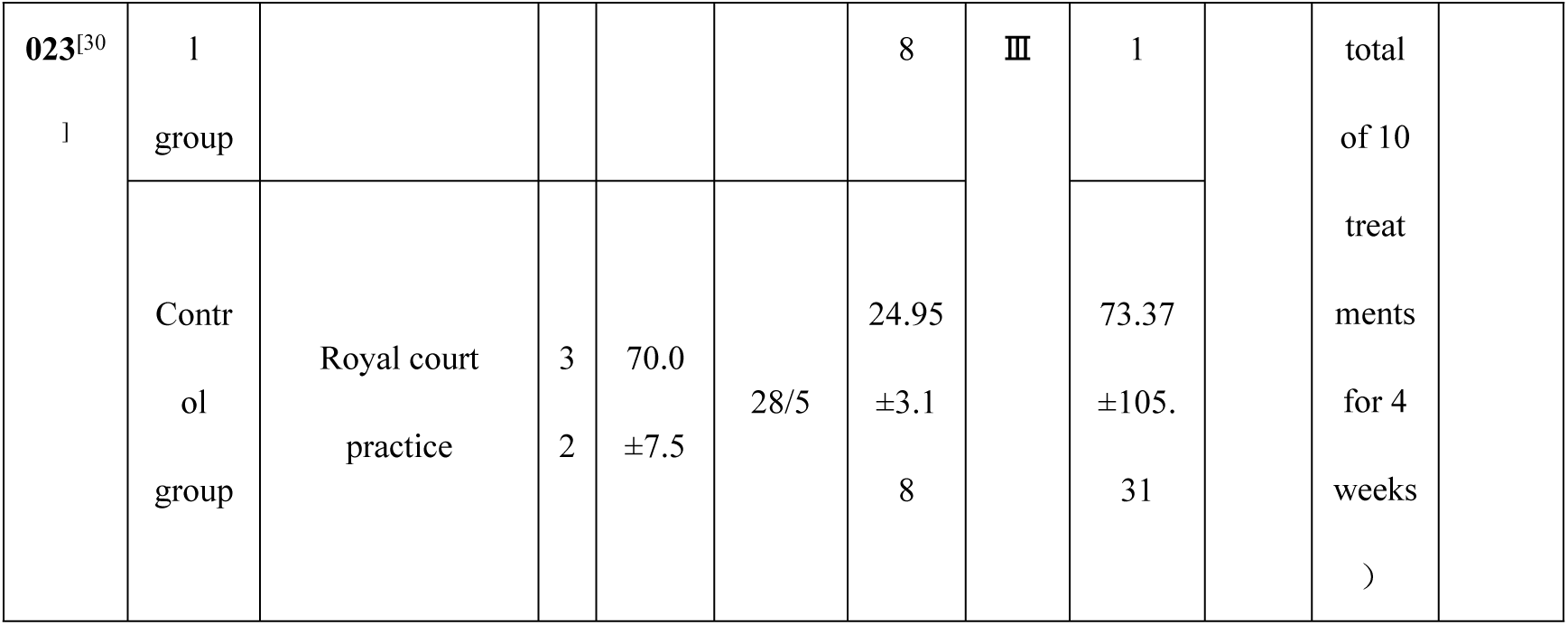
Characteristics of the included studies.

### 3.3 Quality assessment of the included literature

The overall methodological quality of the eight studies included in this meta-analysis was considered to be moderate. All studies utilized a randomized design for group allocation. However, two studies did not provide explicit reporting on the randomization procedure. Among the included studies, various allocation methods were employed: one used a lottery, another used a coin toss, two utilized an envelope system, one employed simple computer-generated randomization, and one used a random number table. Allocation concealment was explicitly mentioned in only six studies. While blinding of patients and outcome assessors was implemented in one study, assessors alone were blinded in four studies, with three studies lacking information on blinding. Detailed summaries of bias risk are depicted in Figure 2A and Figure 2B.

**Figure 2.**
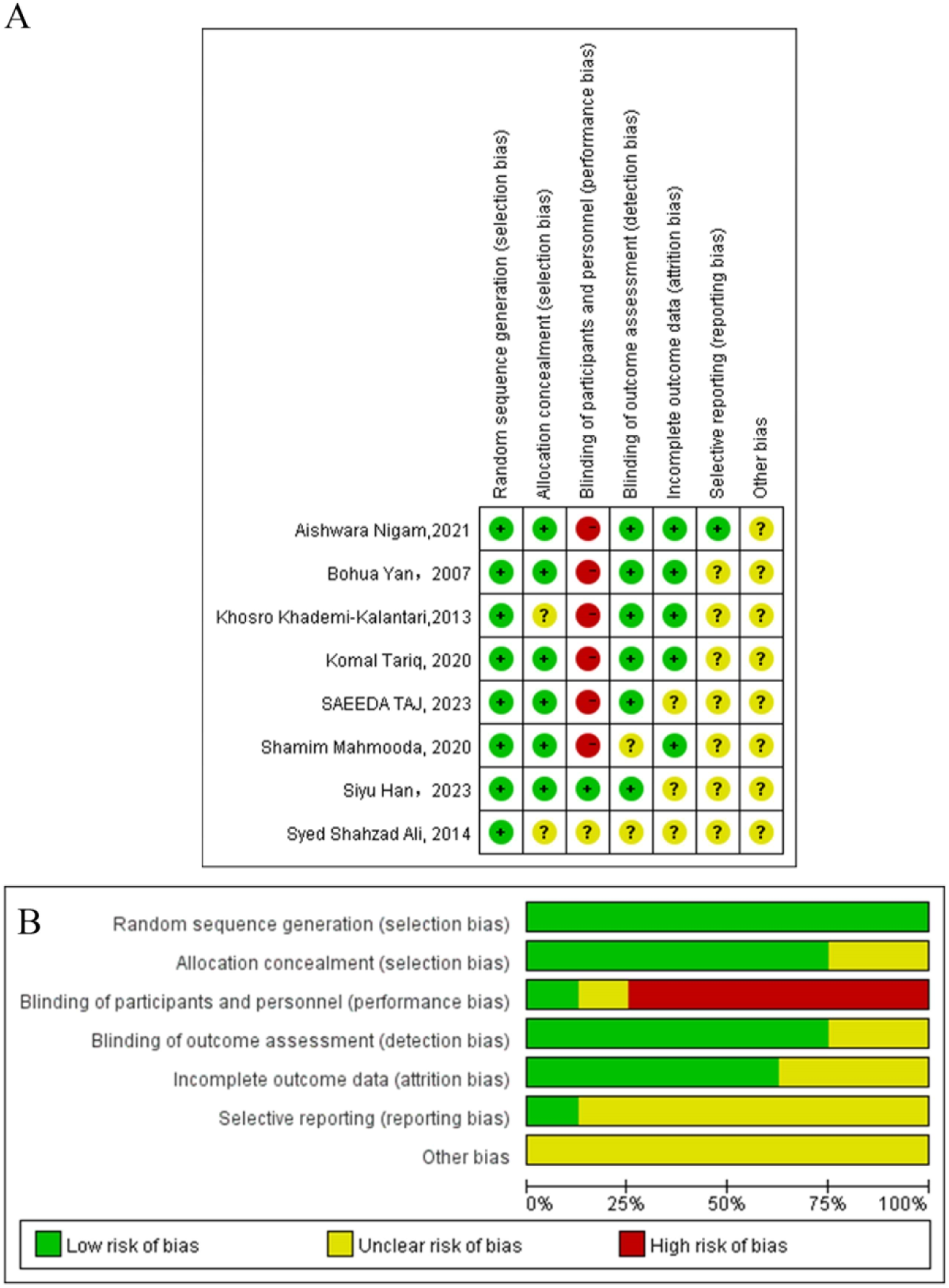
Risk of bias summary and graphs (A): summary of risk of bias, and (B) risk of bias graph.

### 3.4 Results of meta-analysis

#### 3.4.1 VAS

A total of six studies^[24-29]^reported pain scores in patients with KOA after arthrotomy interventions, with four^[25, 27-29]^ using the VAS to report pain intensity and two^[24, 26]^ using the Numeric Pain Rating Scale (NRPS) to report pain intensity. Given that the NRPS and the VAS are used to represent the subjective pain intensity of patients, the NRPS is a segmented numeric version of the VAS, and both use a horizontal line to assess pain levels. Therefore, these two scales can be considered identical.

In the six studies, a total of 159 cases in the experimental group and 160 cases in the control group were included. The test for heterogeneity was P < 0.00001, I^2^ = 90%. Due to the high heterogeneity of the results, we performed subgroup analyses based on different treatment periods, in which four studies with a treatment period of 2 weeks (SMD=-1.98, 95% CI [-3.51, -0.45] Z=2.54 P=0.01) showed a high degree of heterogeneity (I^2^=94%, P<0.00001). Conversely, the other two studies with a treatment period of 4–6 weeks (SMD=- 1.28, 95% CI [-1.74, -0.85] Z=5.43 P<0.0001) had lower heterogeneity (I^2^=35%, P=0.22) (Figure 3A). Sensitivity analysis revealed that Bohua, Yan, 2006 was not in the 95% CI [-1.47, -0.97], suggesting that the result was not robust (Figure 3C). However, when we attempted to exclude Yan BH, 2006, the heterogeneity among studies remained high and did not affect the results of the meta-analysis (Figure 3B).

**Figure 3.**
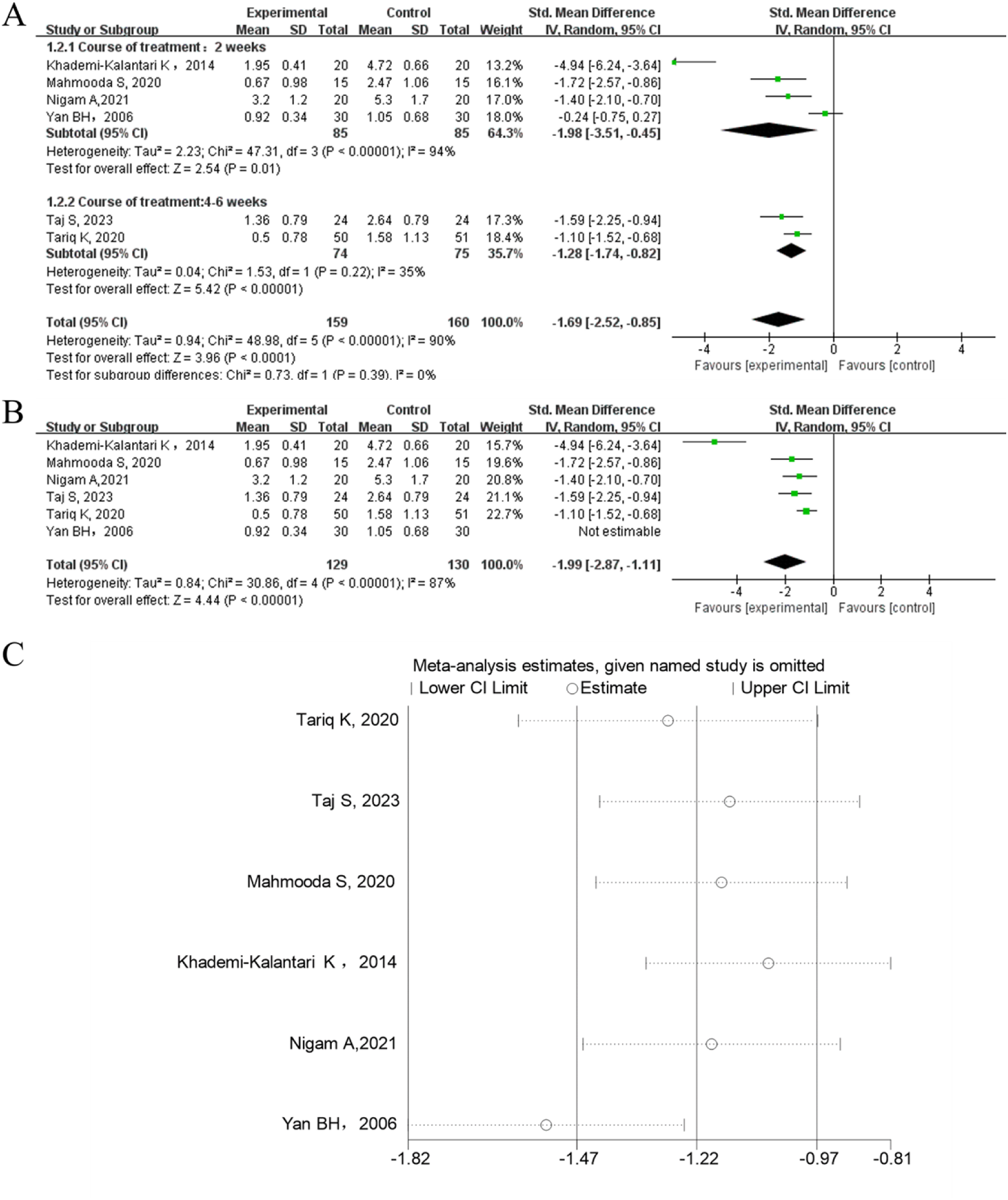
(A) Forest plot for VAS. (B) Forest plot after removing Yan BH, 2007. (C) Plot of sensitivity analysis.

Accordingly, we combined the data using a random-effect model. Meta-analysis showed that SMD = -1.69, 95% CI [-1.74, -0.82] Z = 3.96 P < 0.0001 (Figure 3A). This finding suggested that joint-mobilization technique was effective in improving the level of knee pain in patients with KOA compared with the control group.

#### 3.4.2 WOMAC

A total of five^[24-26, 30, 31]^ studies reported WOMAC scores, with a total of 136 in the experimental group and 137 in the control group included. The test for heterogeneity was P < 0.00001 (I^2^ = 93%) due to the high heterogeneity of results. We conducted subgroup analyses according to the different age groups of the patients. One study^[24]^ with patients aged less than 50 years (SMD=1.15, 95% CI [0.70, 1.60]) had better efficacy in the control group than in the experimental group (Z=5.04, P<0.00001). Three studies^[25, 26, 31]^ conducted on individuals aged 50–60 years (SMD=-1.00, 95% CI [-1.58, -0.43]) had moderate heterogeneity (I^2^= 54%, P=0.11), with better efficacy in the experimental group than in the control group (Z=3.40, P=0.0007). One study^[30]^ on individuals aged >60 years (SMD=-0.05, 95% CI [-0.54, 0.45]) had no significant difference between the control and experimental groups (Z=0.18, P=0.85). Using a random-effect model to combine the data, results showed that SMD=-0.36,95% CI [- 1.31, -0.59], and no significant difference existed between the experimental and control groups (Z=0.75, P=0.45) (Figure 4A). Sensitivity analysis revealed that this result was not robust (Figure 4C).

**Figure 4.**
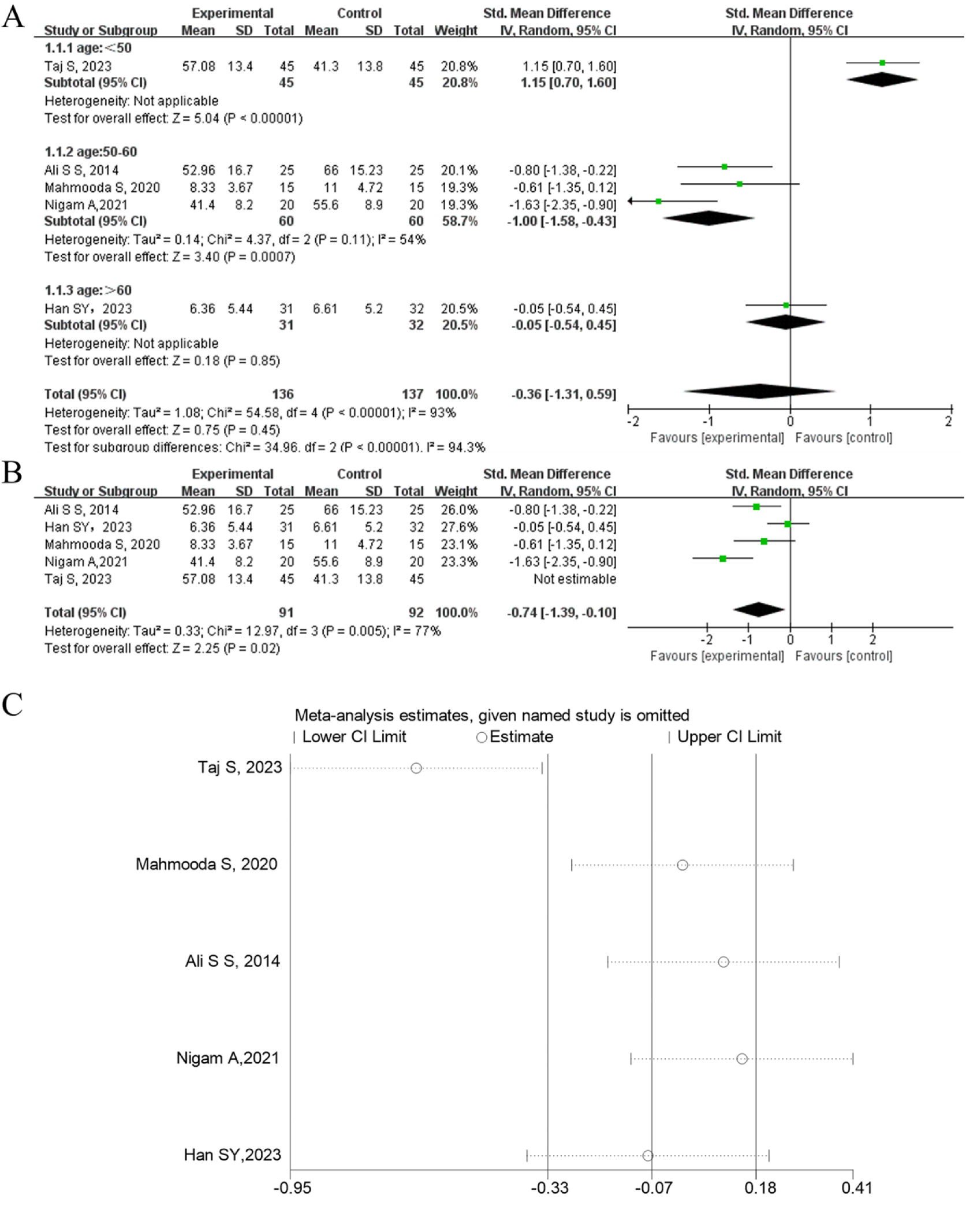
(A) Forest plot for WOMAC. (B) Forest plot after removing Taj S, 2023. (C) Plot of sensitivity analysis.

We found that a new SMD of -0.74 (95% CI [-1.39, -0.10]) was generated when Taj S, 2023^[24]^ was excluded, which may be due to the fact that Taj S, 2023 included KOA patients with pain combined with limited knee mobility, inconsistent with the baseline level of patients included in other studies. This outcome indicates a notable improvement in knee function performance among patients with KOA in the experimental group compared to the control group. (Z=2.25, P=0.02) (Figure 4B).

#### 3.4.3 Knee flexion angle

A total of three groups^[24, 26, 28]^ of studies reported the knee flexion angle after the intervention, with a total of 80 cases included in the experimental group and 80 cases in the control group. The result of the heterogeneity test was P < 0.00001 I^2^ = 93%. Due to the high heterogeneity of the result, we performed subgroup analyses based on different treatment periods. Two of the studies^[26, 28]^ had a treatment period of 2 weeks (SMD=2.30, 95% CI [0.98, 3.62]) with high heterogeneity (I^2^=78%, P=0.03). The efficacy of the control group was better than that of the experimental group (Z=3.41, P=0.0006), and one study^[24]^ had a treatment period of 6 weeks (SMD=-1.97, 95% CI [-2.48, -1.47]) The experimental group was superior to the control group (Z=7.62, P<0.00001). Using a random-effect model to combine the data, results showed that SMD=0.87,95% CI [-2.33,4.07], and no significant difference existed between the experimental and control groups in terms of improving the knee flexion angle (Z=0.53, P=0.60)(Figure 5A). Sensitivity analysis showed it was not robust (Figure 5C).

**Figure 5.**
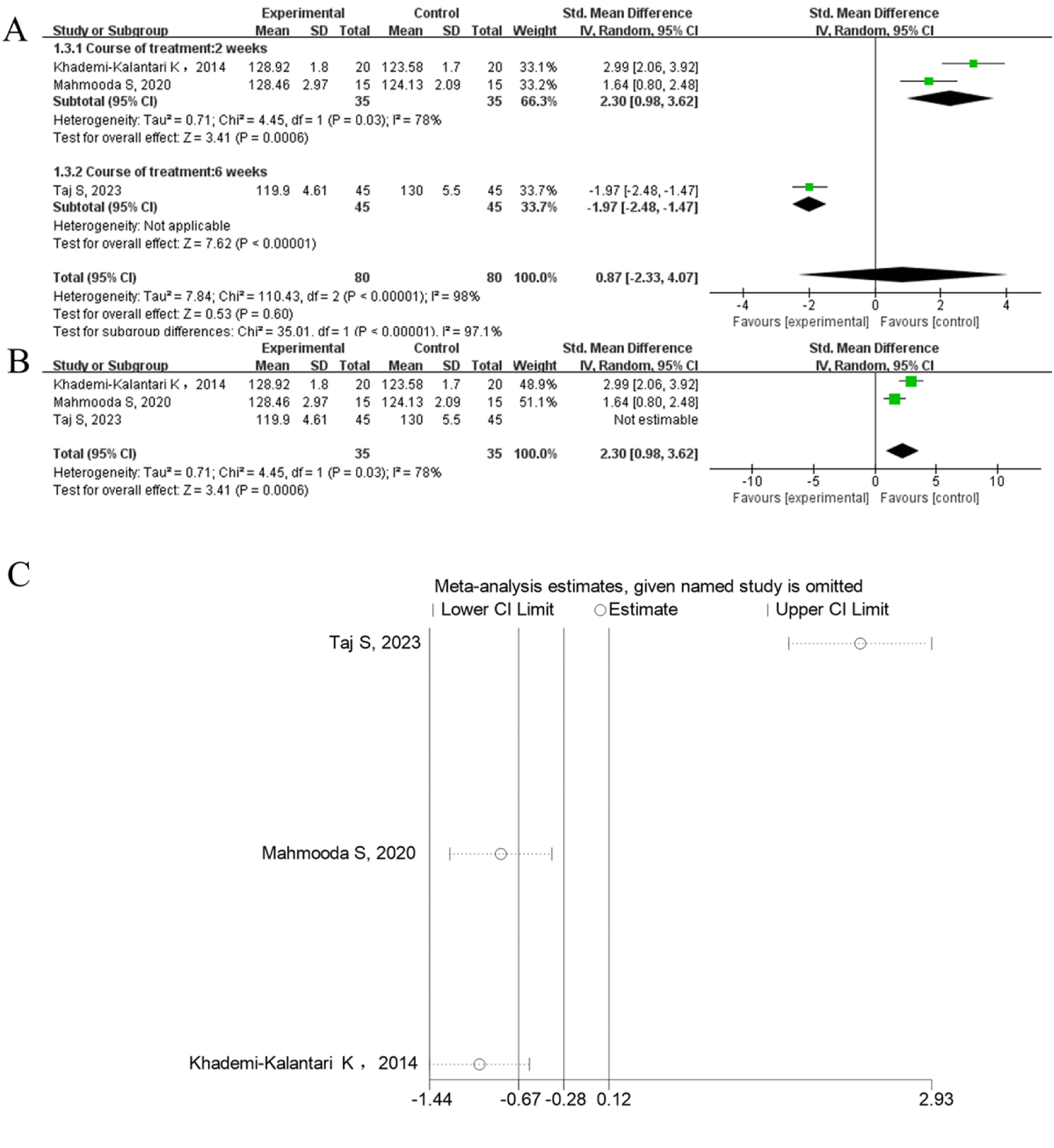
(A) Forest plot for flexion angle. (B) Forest plot after removing Taj S, 2023. (C) Plot of sensitivity analysis.

Similarly, when we attempted to exclude Taj S, 2023^[24]^, we found a new SMD of 2.3 (95% CI [0.98, 3.62], which may remain due to the fact that the patients included in Taj S, 2023 were not consistent with the other studies. This result showed that the intervention was less efficacious in improving knee flexion angle compared with the control group, and the difference was statistically significant (Z=3.41, P=0.00006) (Figure 5B).

#### 3.4.4 Knee-extension angle

A total of two trials^[24, 28]^reported the knee-joint extension angle after the intervention, which included 65 cases in the experimental group and 65 in the control group. No heterogeneity existed between the two groups (P=0.73, I^2^=0%), so we use a fixed-effect model for the meta-analysis. Results showed that SMD=1.79, 95% CI [1.38, 2.20], and Z=8.54,P<0.00001. This finding indicated that the efficacy of the joint-mobilization technique was lower in improving the knee-extension angle compared with the control group, and the difference was statistically significant (Figure 6A). Sensitivity analysis indicated that this result was robust (Figure 6B).

**Figure 6.**
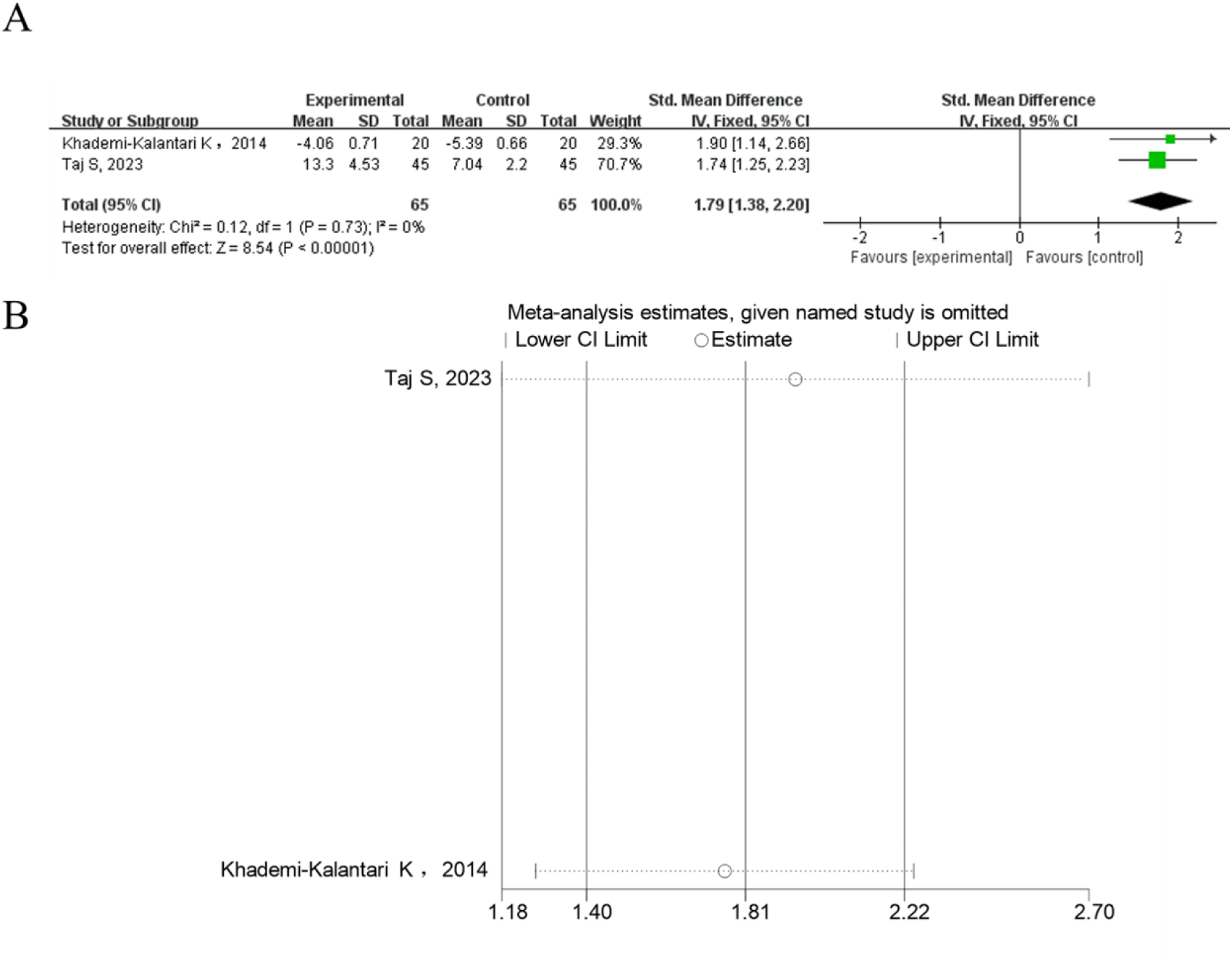
(A) Forest plot for extension angle. (B) Plot of sensitivity analysis.

#### 3.4.5 Publication bias

Egger test showed it was no significant publication bias in VAS, WOMAC (P > 0.05), but a significant publication bias existed in knee flexion angle (P = 0.037); The extension angle had only two trails and was not evaluated for publication bias. Figure 7.

**Figure 7.**
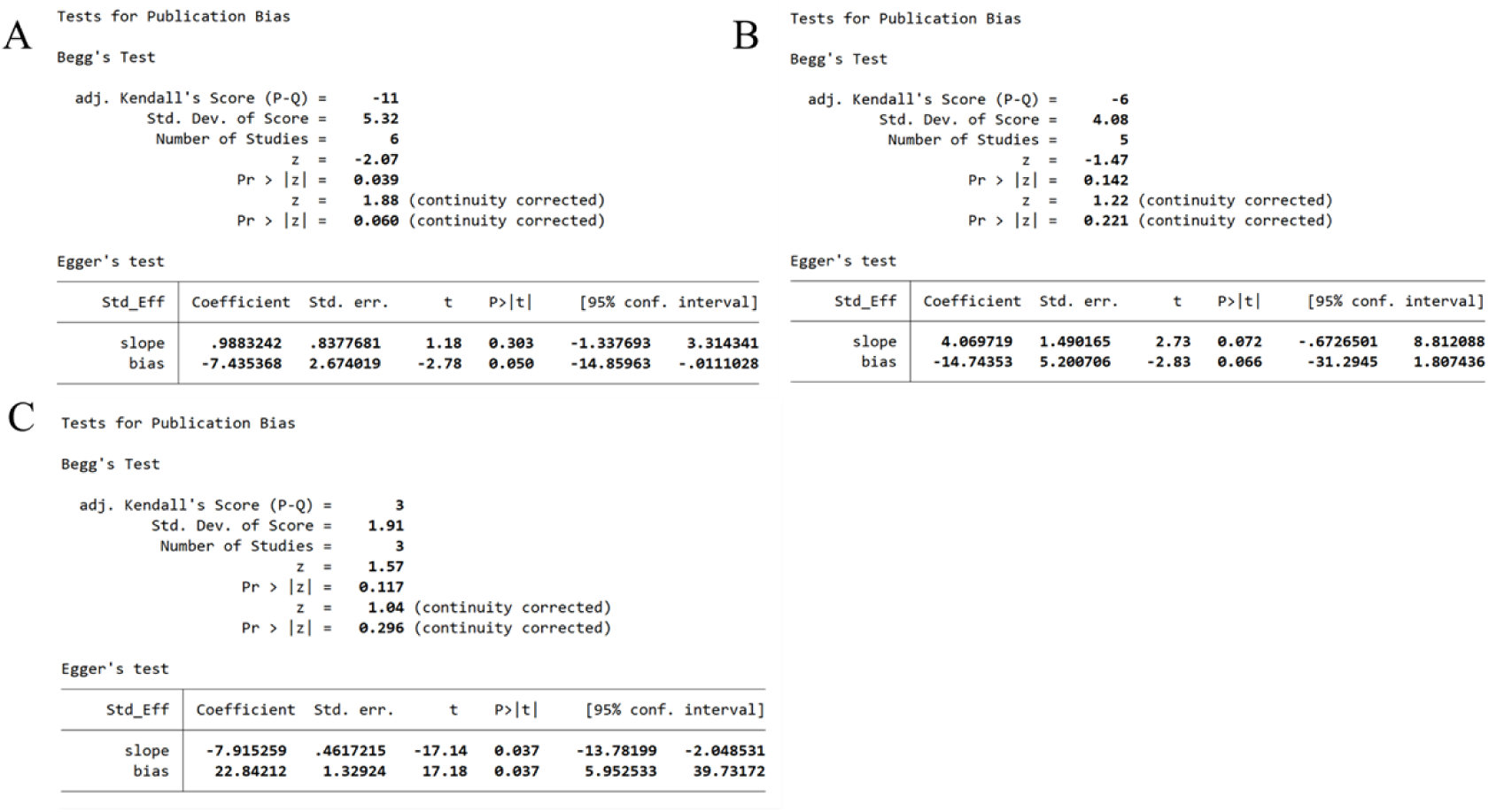
Plots of publication bias: (A) VAS, (B) WOMAC, (C) flexion angle.

## 4 Discussion

This meta-analysis aimed to evaluate the effect of joint-mobilization technique on knee pain, function, and mobility in patients with KOA. The pooled analysis included eight eligible randomized controlled trials from several countries. Meta-analysis results revealed that joint-mobilization technique can significantly improve knee pain levels and dysfunction compared with controls, but it was not preferred in improving knee-joint mobility. Thus, joint-mobilization technique can be serve as an adjunct treatments in KOA patients with severe pain or dysfunction.

Six articles^[24-29]^ included in this study were meta-analyzed using knee pain level as an evaluation index. We restricted the intervention periods to 2–6 weeks, but significant variability remained among the studies. This finding suggested that the intervention duration significantly influenced the joint-mobilization efficacy. We used different treatment periods as the basis for subgroup analyses, but significant heterogeneity remained between the 4 studies with 2-week duration. Then, we tried to analyze the effects of gender, age, duration of treatment, mode of intervention in the control group or KOA grading. Unfortunately, they were not the cause of high heterogeneity. We then found through sensitivity analysis that heterogeneity remained high after excluding Yan BH, 2006^[29]^. We reviewed the original literature and found that the study included KOA patients with a KL classification of 0. This finding may be the reason why the Yan BH, 2006 95% CI did not fall within the overall CI when the sensitivity analysis was performed. It led to an unrobust result, but it still did not explain the high heterogeneity. Some of the articles did not provide the duration of the patient’s KOA disease, the frequency of interventions, and the type of mobilization chosen, so we cannot determine whether these factors were a cause of heterogeneity. Moreover, joint-mobilization technique is a form of manipulative intervention performed exclusively by a doctor or therapist. The strength, direction, and type of application are all subjective, but all of these factors affect clinical efficacy and assessment, which may also contribute to the high heterogeneity. Egger test showed no significant publication bias, suggesting that joint-mobilization technique can be an effective manipulative therapeutic intervention to alleviate the level of knee pain in patients with KOA.

In the WOMAC evaluation analysis, we used different age groups of patients as the basis for subgroup analyses. Results showed that joint-mobilization technique performed better than the control group only in patients aged 50–60 years. Sensitivity analyses revealed inconsistent results when Taj S, 2023^[24]^ was excluded, which may be due to the fact that the patients selected at Taj S, 2023 were all patients with KOA with pain combined with limited knee mobility, resulting in a new result due to inconsistencies in the baseline level with other studies. The Egger test showed no significant publication bias. Based on these results, we can conclude that arthroplasty may be more suitable for people aged 50–60 years to improve knee function. However, this result does not provide sufficient evidence due to the few articles included in each age group.

In terms of knee-joint mobility, our meta-analyses all showed that joint-mobilization technique was not superior to the control group in improving knee flexion and extension angles. This finding was inconsistent with what we expected. We speculated that it may be due to the fact that most of the KOA patient’s visit narrative was to improve joint pain, so the technique applied by the therapist was primarily for knee pain relief, such as level I or II manipulation, rather than level III or IV manipulation for improving mobility. This result was consistent with that reported by Ling-Ling Li^[23]^. Our sensitivity analyses showed that the results for flexion was unrobust, but the extension was robust. Additionally, when Taj S, 2023 was excluded, flexion showed a different result, which may be due to the fact that the patients selected at Taj S, 2023 were all KOA patients with pain combined with limited knee mobility, thereby generating a new result. Egger test indicated a significant publication bias in the knee flexion angle, which may be due to the small number of articles included or the low quality of the articles. Our results suggested that none of the joint-mobilization technique was better than the control group in improving range of motion, but this result may be unreliable because only two to three papers reported knee mobility, which may have led to biased and low-quality results. Thus, more literature data are needed to delve deeper into this conclusion.

Although this meta-analysis was strictly based on inclusion and exclusion criteria to screen the relevant literature, this study had some limitations. First, the sample sizes of the included studies were all less than 100 people, which may have affected the quality and accuracy of the results. Second, the methodological limitations of the study, due to the specificity of joint mobilization technique, rendered impossible the blinding of patients and therapists at the same time, leading to a high risk of bias. Although blinding the patients was not feasible, efforts were made to minimize other sources of bias to mitigate overall study bias. Additionally, the limited number of included articles, particularly concerning knee flexion and extension angles, likely influenced the bias of the findings, thus impacting the overall results. Fourth, KOA is a global disease and most of the articles included in this study were conducted in Asia, possibly misrepresenting the global population. Fifth, the outcomes are limited, including only WOMAC, VAS, knee flexion and extension angles; thus, objective markers such as laboratory diagnostics and imaging tests are lacking. Sixth, most of the experimental group interventions in this study were combined with other interventions such as physical factor therapy and exercise training. We were unable to exclude the interference of other interventions, which led to a bias in the overall results. Hence more high-quality clinical randomized controlled trials with joint mobilization alone as the only intervention are needed. Seventh, the intervention effects were observed only in the 2–6 week treatment period, and no study was made on the long-term effects. Finally, the optimal intervention time, frequency, and intensity of the treatment were not explained. In conclusion, we need more high-quality clinical studies with large samples, multiple intervention doses, and multicenters to explore clearer and more reliable treatments and thus improve clinical decision-making for KOA treatment.

Joint-mobilization technique is a form of manipulative intervention, whereas current clinical trials still suffer from methodological shortcomings and potentially biased findings. In future clinical trials, more large-sample, multicenter randomized controlled trials are needed. KOA, as a long-term chronic disease, requires more focus on the duration and long-term effects of the intervention. More standard protocols for different ages or severity levels also need to be developed to provide effective treatment options for KOA patients.

## 5 Conclusion

Joint-mobilization technique is effective in improving knee pain and function in patients with KOA compared with the control group. No difference was found in knee mobility. However, due to each result’s high heterogeneity and the influenced of literature quality, this conclusion needs more high-quality evidence to support it.

## Data Availability

All relevant data are within the manuscript and its Supporting Information files.

## Abbreviations

KOA: Knee Osteoarthritis
WOMAC: Western Ontario and McMaster Universities Arthritis Index
CI: confidence interval
SMD: standard mean difference.

